# A genomic perspective on the near-term impact of doxycycline post-exposure prophylaxis on *Neisseria gonorrhoeae* antimicrobial resistance

**DOI:** 10.1101/2023.03.14.23287223

**Authors:** Tatum D. Mortimer, Yonatan H. Grad

## Abstract

Post-exposure prophylaxis with doxycycline (doxyPEP) is being introduced to prevent bacterial sexually transmitted infections (STIs). Pre-existing tetracycline resistance in *Neisseria gonorrhoeae* limits doxyPEP effectiveness against gonorrhea, and selection for tetracycline resistant lineages may influence prevalence of resistance to other antimicrobials via selection for multi-drug resistant strains. Using genomic and antimicrobial susceptibility data from 5,644 clinical isolates of *N. gonorrhoeae*, we assessed the near-term impact of doxyPEP on *N. gonorrhoeae* antimicrobial resistance. We found that the impact on antimicrobial resistance is likely to be influenced by the strength of selection for plasmid-encoded and chromosomally-encoded tetracycline resistance, as isolates with high-level, plasmid-encoded resistance had lower minimum inhibitory concentrations to other antimicrobials compared to isolates with low-level tetracycline resistance. The impact of doxyPEP may differ across demographic groups and geographic regions within the United States due to variation in pre-existing tetracycline resistance.

---

Post-exposure prophylaxis with doxycycline (doxyPEP) has been shown to decrease rates of bacterial sexually transmitted infections (STIs) in clinical trials,^1–3^ and doxyPEP is recommended by some public health departments in the United States. Official guidelines from the US Centers for Disease Control and Prevention (CDC) and other public health institutions are pending.

A concern around doxyPEP is its impact on antibiotic resistance. Pre-existing tetracycline resistance in the *Neisseria gonorrhoeae* population resulted in lower doxyPEP effectiveness against gonorrhea than against syphilis and chlamydia^2^ and reflects tetracycline and multidrug resistant strains of *N. gonorrhoeae* that may be selected for by doxyPEP. Given this concern, we evaluated the potential near-term impacts of doxyPEP on antimicrobial resistance in *N. gonorrhoeae* using whole genome sequencing (WGS) data and minimum inhibitory concentrations (MICs) from a global collection of 5,644 *N. gonorrhoeae* isolates (Supplementary Tables 1-2), including 1,041 isolates from 2018 collected and sequenced by CDC’s Gonococcal Isolate Surveillance Program (GISP)^4^.

Interpretative breakpoints for *N. gonorrhoeae* susceptibility and resistance to doxycycline have not been defined; however, treatment failures have been observed when isolates have doxycycline MICs ≥ 1 µg/ml,^5^ and doxycycline MICs correlate with tetracycline MICs.^6^ Tetracycline resistance can be mediated by plasmid-encoded *tetM*, which confers high-level resistance, and chromosomally-encoded mutations in *rpsJ, porB*, and the *mtr* operon.^7^ We excluded isolates that had tetracycline MICs that most likely represented reporting errors (Supplementary Text).

We found that co-resistance to other antimicrobials was most common in isolates with chromosomally-encoded tetracycline resistance (Figure 1). Ceftriaxone, azithromycin, and ciprofloxacin MICs were significantly higher in isolates with tetracycline MICs of 2-8 µg/ml compared to isolates not resistant to tetracycline and to isolates with high-level tetracycline resistance (p < 0.0001, Mann-Whitney test). Ceftriaxone reduced susceptibility (MIC ≥ 0.125 µg/mL) was rare in GISP isolates (n=2), but the global data suggested that reduced susceptibility is most often acquired by strains with chromosomally-mediated tetracycline resistance, including strains encoding the *penA* 60 allele, which confers ceftriaxone reduced susceptibility.^8^ Azithromycin resistance appeared in 23.5% (56/238) of GISP isolates resistant to tetracycline; all that were tetracycline and azithromycin co-resistant had chromosomally encoded tetracycline resistance. Ciprofloxacin resistance appeared in 33.6% (80/238) of GISP isolates resistant to tetracycline; however, only 12.9% (13/101) of isolates with *tetM* were also ciprofloxacin resistant.

**Figure 1.**
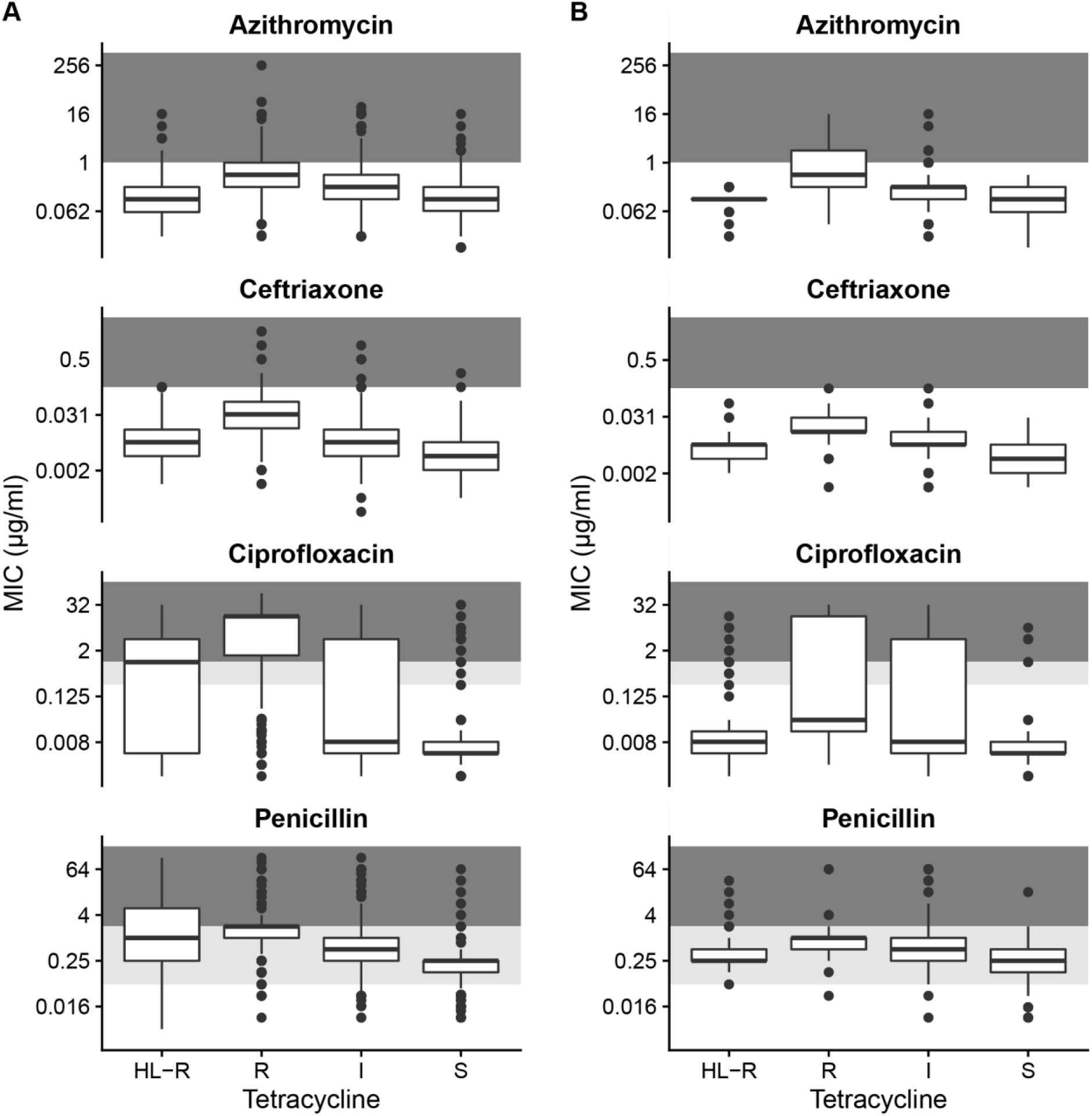
Co-resistance with other antimicrobials is highest among isolates with chromosomally-encoded resistance to tetracyclines. Isolates were classified as susceptible (MIC ≤ 0.25 µg/mL), intermediate (0.25 < MIC < 2 µg/mL), resistant (2 ≤ MIC ≤ 8 µg/mL), or high-level resistant (MIC > 8 µg/mL) in 5,644 global *N. gonorrhoeae* isolates (**A**) and 1,041 isolates collected in the United States in 2018 (**B**). Background shading corresponds to susceptible (white), intermediate (light gray), and resistant/non-susceptible (dark gray) MICs for each antimicrobial.

In the United States, doxyPEP is primarily recommended for men who have sex with men (MSM) and transgender persons who have sex with men. The distribution of isolates classified as susceptible (MIC ≤ 0.25 µg/mL), intermediate (0.25 < MIC < 2 µg/mL), resistant (2 ≤ MIC ≤ 8 µg/mL), or high-level resistant (MIC > 8 µg/mL) significantly differed between MSM and men who have sex with women (MSW) (Supplementary Table 3, p < 0.0001, χ^2^ test). While the proportion of isolates with tetracycline resistance was similar between MSM (26.8%, 91/340) and MSW (21.3%, 125/587), tetracycline susceptibility was more common among MSW (23.0%, 135/587) compared to MSM (10.3% 35/340). Among MSM, the majority of isolates (62.9%, 214/340) had intermediate tetracycline MICs. In addition to selection for resistant lineages in MSM populations, this large population of *N. gonorrhoeae* with intermediate MICs may represent a reservoir for rapid evolution of resistance. However, the evolutionary pathways and barriers to doxycycline resistance and their variation by genomic background in *N. gonorrhoeae* are unknown.

The composition of *N. gonorrhoeae* isolates was not homogeneous across the United States, and the percentage of tetracycline resistant isolates collected across HHS regions ranged from 17.4% - 52.0% (Supplementary Figure 1). Mechanisms of resistance also varied, and high-level resistance was not evenly distributed across geographic regions, with the lowest proportion of isolates with high-level resistance in HHS regions from the east coast and southeastern United States (1.7% - 4.4%). This indicates that doxyPEP will likely have regionally varying effectiveness and impact on circulating *N. gonorrhoeae*.

In our study, the genomic data representing the *N. gonorrhoeae* population in the United States were from isolates collected in 2018, and recent changes in treatment guidelines^9^ and introductions of resistant lineages, such as isolates encoding *penA* 60,^10^ may have shifted the landscape of antimicrobial resistance in the United States. The number of genomes meeting quality thresholds from the southeastern United States was low (Supplementary Figure 1), so these data may not accurately capture lineages transmitting in this region. We emphasize that the long-term impacts of doxyPEP on the *N. gonorrhoeae* population in the United States are unknown, as lineages may acquire new resistance alleles and introduction of lineages from other geographic regions is possible.^11,12^ For example, a recent study found that strains encoding *tetM* are highly prevalent among women in Kenya,^13^ and phylogenetic analysis of our global dataset identified several lineages with high-level tetracycline resistance not represented among GISP isolates (Supplementary Figure 2).

The near-term impact of doxyPEP on *N. gonorrhoeae* antimicrobial resistance and circulating lineages will be influenced by the strength of selection for tetracycline resistance. If doxyPEP use selects for all tetracycline resistant lineages, there is potential for increased resistance to other antimicrobials as well. However, if doxyPEP use primarily selects for lineages with *tetM*-mediated resistance, we may observe a temporary decline in resistance to other antimicrobials due to relatively less co-resistance in these lineages. Timely genomic surveillance, with a focus on doxyPEP users, is needed to distinguish between these outcomes.

## Data Availability

All data produced are available at https://github.com/gradlab/doxyPEP_genomics.

## Acknowledgments

This work was funded by NIAID R01 AI132606 and R01 AI153521. The funding sources had no role in the writing of the manuscript or the decision to submit it for publication.

## Author contributions

TDM performed data curation and analysis. YHG supervised and managed the study. TDM and YHG wrote the manuscript, had full access the data reported, and were responsible for the decision to submit the manuscript for publication.

