## Supplementary Text for "A genomic perspective on the near-term impact of doxycycline post-exposure prophylaxis on *Neisseria gonorrhoeae* antimicrobial resistance"

### Supplementary Material

### Table of Contents

Methods. 2
Dataset. 2
Genome Assembly. 2
Quality Control of Genomic Data. 2
Phylogenetic Analysis. 2
Statistical analysis. 2
Data availability. 2
Tetracycline resistance genotypes and phenotypes. 2
Supplementary Table 1. Genomic datasets with tetracycline MICs. 4
Supplementary Table 2. Quality control metrics. 4
Supplementary Table 3. Tetracycline susceptibility among sexual behavior groups in the United States, 2018. 4
Supplementary Figure 1. Tetracycline susceptibility across geographic regions. 5
Supplementary Figure 2. Phylogeny of 5,445 isolates with tetracycline MICs 6
References. 7

**Methods.**

*Dataset.* Whole genome sequencing data and minimum inhibitory concentrations were collected from previously published *N. gonorrhoeae* genomic studies (Supplementary Table 1). Isolates were included if the tetracycline MIC was reported with sufficient precision to categorize isolates as susceptible, intermediate, resistant, or high-level resistant.

*Genome Assembly.* Whole genomes sequencing data was assembled as previously described.[1] Briefly, we used SPAdes v 3.12.0[2] for *de novo* assembly; the –careful flag was used to correct assemblies, and contigs were removed if coverage was less than 10X or contig length was less than 500 nucleotides. Reads were additionally mapped to the NCCP11945 reference genome (NC_011035.1) using BWA-MEM v 0.7.17[3], and variants were called using Pilon v 1.23[4] with a minimum mapping quality of 20 and 10X minimum coverage.

*Quality Control of Genomic Data.* We included isolates with genomes meeting the following quality control filters: the number of contigs was less than 500, the assembly length and number of annotated genes was expected for *N. gonorrhoeae,* coverage was greater than 30X, at least 80% of reads mapped to the *N. gonorrhoeae* reference genomes, and fewer than 12% of sites in the *N. gonorrhoeae* reference genome were unable to be confidently called using our assembly pipeline. Quality control metrics for included genomes is summarized in Supplementary Table 2. Additionally, we required that the contig encoding *tetM* was greater than 10,000 nucleotides in length and at least 15X coverage.

*Phylogenetic Analysis.* We used pseudogenomes derived from reference mapping and variant calls for phylogenetic analysis; we required that 90% of reads supported an allele to include the position in an isolate's pseudogenome. We used Gubbins v 2.4.1[5] to identify recombinant regions and reconstruct the phylogeny.

*Statistical analysis.* All statistical analysis was performed in R v 4.1.3.[6] Significance of differences between MICs to ceftriaxone, azithromycin, ciprofloxacin, and penicillin was assessed using a Mann-Whitney test. Significance of differences of susceptible, intermediate, resistant, and high-level resistant isolates among demographic groups was assessed using a $\chi^2$ test.

*Data availability.* Data and code are available at https://github.com/gradlab/doxyPEP_genomics

**Tetracycline resistance genotypes and phenotypes.**

In this dataset, 97.3% (1018/1046) of isolates with high-level tetracycline resistance (MIC > 8 µg/ml) encoded *tetM*. Tetracycline MICs ranging from 1 – 8 µg/ml were primarily associated with chromosomally-encoded resistance mutations, and only 6.2% (67/1085) of isolates encoding *tetM* had a reported MIC ≤ 8 µg/ml.

Given the expected error of up to two doubling dilutions in MIC measurements,[7] we investigated isolates encoding *tetM* with MICs < 4 µg/mL, representing unexplained increased tetracycline susceptibility. We hypothesized that isolates encoding a genetic variant that significantly impacted MICs would cluster with isolates with similarly low MICs, whereas isolates with errors in MIC reporting would likely appear as singletons among isolates with MICs consistent with their genotype. In keeping with the hypothesis that these were likely MIC reporting errors, the isolates with unexplained increased low MICs (n = 22, 2.0% of isolates with *tetM*) did not cluster in the phylogeny, and isolates with unexplained high-level resistance (n = 28) were singletons (Supplementary Figure 1). A total of 50 isolates, including 11 GISP 2018 isolates, were excluded from further analysis.

**Supplementary Table 1. Genomic datasets with tetracycline MICs.**

| Reference | Number of Isolates passing QC |
|---|---|
| Alfsnes et al. 2020[8] | 841 |
| Bristow and Mortimer et al. 2023[9] | 443 |
| Demczuk et al. 2015[10] | 89 |
| Eyre et al. 2017[11] | 231 |
| Ezewudo et al. 2015[12] | 51 |
| Golparian et al. 2020[13] | 665 |
| Golparian et al. 2022[14] | 61 |
| Grad et al. 2016[15] | 1093 |
| Lan et al. 2020[16] | 226 |
| Lee et al. 2018[17] | 392 |
| Pinto et al. 2020[18] | 134 |
| Reimche et al. 2021[19] | 1040 |
| Sánchez-Busó et al. 2018[20] | 378 |

**Supplementary Table 2. Quality control metrics.**

| Metric | Median | Minimum | Maximum |
|---|---|---|---|
| Number of contigs | 98 | 59 | 424 |
| Assembly Length | 2132963 | 1966448 | 2258145 |
| Number of annotated genes | 2090 | 1917 | 2285 |
| Coverage | 123 | 32 | 1382 |
| Percentage of reads mapped to reference | 93.3 | 80.0 | 99.7 |
| Percentage of reference genome with missing or ambiguous calls | 6.7 | 3.2 | 12.0 |

**Supplementary Table 3. Tetracycline susceptibility among sexual behavior groups in the United States, 2018.** Isolates were collected and sequenced by CDC's Gonococcal Isolate Surveillance Program in 2018.[19]

| Sexual Behavior* | S MIC ≤ 0.25 μg/mL | | I 0.25 < MIC < 2 μg/mL | | R 2 ≤ MIC ≤ 8 μg/mL | | HL-R MIC > 8 μg/mL | |
|---|---|---|---|---|---|---|---|---|
| | Total | Percent | Total | Percent | Total | Percent | Total | Percent |
| MSM | 35 | 10.3% | 214 | 62.9% | 54 | 15.9% | 37 | 10.9% |
| MSW | 135 | 23.0% | 327 | 55.7% | 75 | 12.8% | 50 | 8.5% |
| MSMW | 9 | 13.2% | 41 | 60.3% | 10 | 14.7% | 8 | 11.8% |

* MSM: men who have sex with men, MSW: men who have sex with women, MSMW: men who have sex with men and women

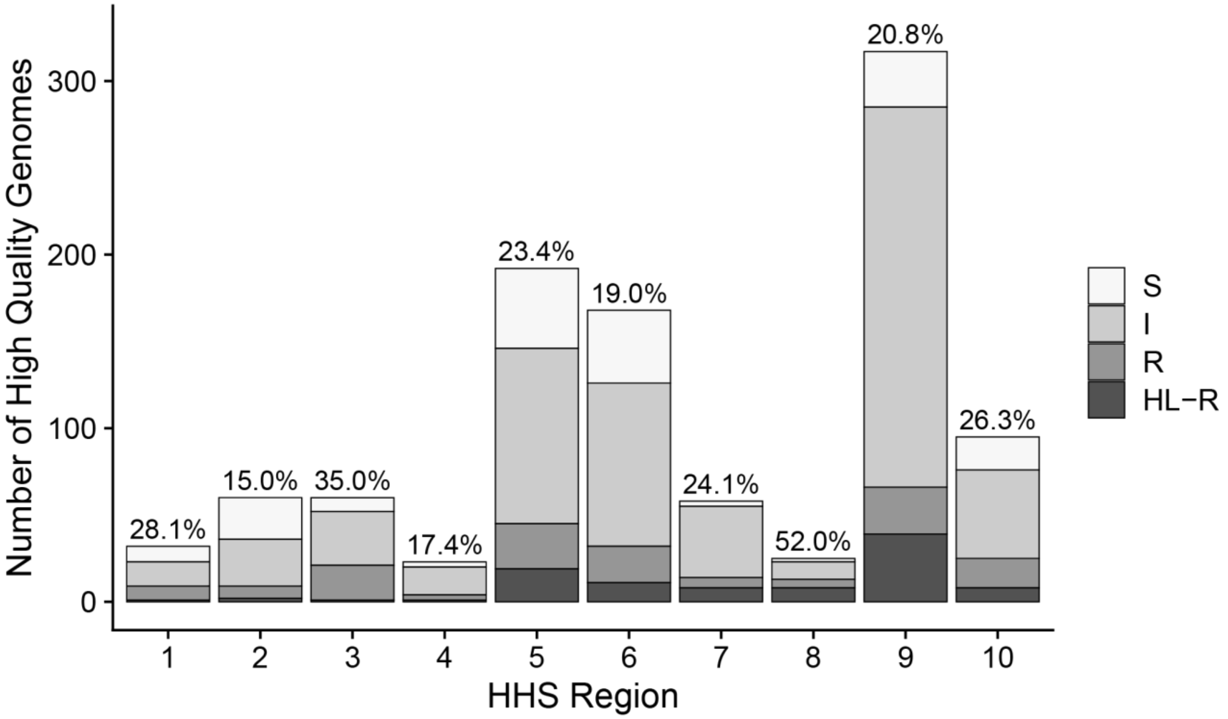


**Supplementary Figure 1. Tetracycline susceptibility across geographic regions.** Bars are shaded based on the number of isolates classified as susceptible (S, MIC ≤ 0.25 μg/mL), intermediate (I, 0.25 < MIC < 2 μg/mL), resistant (R, 2 ≤ MIC ≤ 8 μg/mL), or high-level resistant (HL-R, MIC > 8 μg/mL). Bars are labeled with the percentage of isolates resistant to tetracycline via low- or high-level resistance (MIC ≥ 2 μg/mL) in each HHS region.

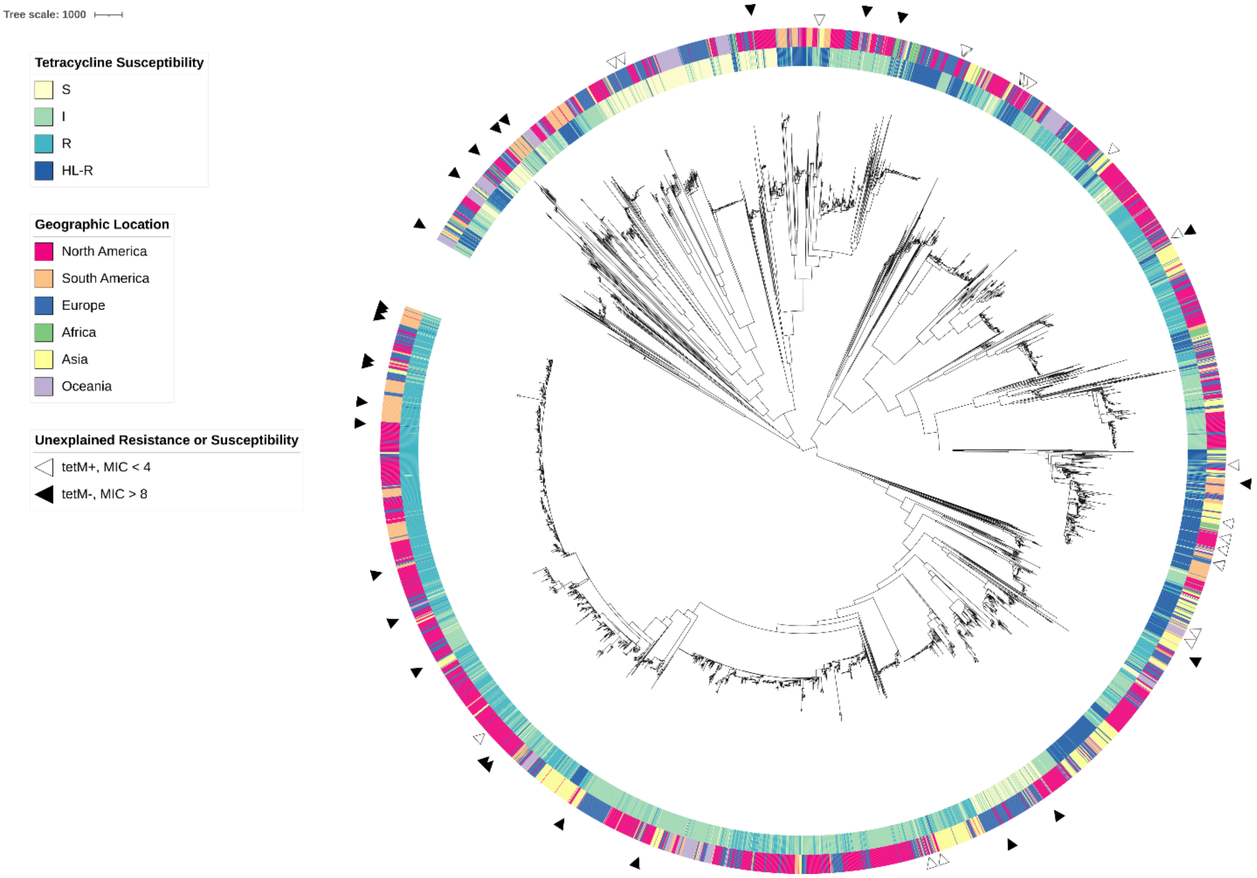


**Supplementary Figure 2. Phylogeny of 5,445 isolates with tetracycline MICs.** The inner ring represents tetracycline susceptibility: susceptible (MIC ≤ 0.25 µg/mL) isolates are yellow, intermediate (0.25 < MIC < 2 µg/mL) isolates are green, resistant (2 ≤ MIC ≤ 8 µg/mL) isolates are teal, and high-level resistant (MIC > 8 µg/mL) isolates are dark blue. The outer ring represents the geographic location from which the isolate was sampled. White arrows indicate isolates encoding *tetM* with unexplained increased susceptibility (MIC < 4). Black arrows indicate isolates that do not encode *tetM* with unexplained high-level resistance (MIC > 8). Additional annotations, including the presence of tetracycline resistance associated *tetM* and *rpsJ* V57M, and *penA* alleles are available at https://itol.embl.de/tree/134174250158349781673367540#.

**References.**


1 Mortimer TD, Zhang JJ, Ma KC, Grad YH. Loci for prediction of penicillin and tetracycline susceptibility in Neisseria gonorrhoeae: a genome-wide association study. *The Lancet Microbe* 2022; **3**: e376–81.

2 Bankevich A, Nurk S, Antipov D, *et al.* SPAdes: A New Genome Assembly Algorithm and Its Applications to Single-Cell Sequencing. *Journal of Computational Biology* 2012; **19**: 455–77.

3 Li H. Aligning sequence reads, clone sequences and assembly contigs with BWA-MEM. *arXiv:13033997 [q-bio]* 2013; published online March 16. http://arxiv.org/abs/1303.3997 (accessed Sept 29, 2015).

4 Walker BJ, Abeel T, Shea T, *et al.* Pilon: An Integrated Tool for Comprehensive Microbial Variant Detection and Genome Assembly Improvement. *PLOS ONE* 2014; **9**: e112963.

5 Croucher NJ, Page AJ, Connor TR, *et al.* Rapid phylogenetic analysis of large samples of recombinant bacterial whole genome sequences using Gubbins. *Nucl Acids Res* 2015; **43**: e15–e15.

6 R Core Team. R: A Language and Environment for Statistical Computing. Vienna, Austria: R Foundation for Statistical Computing, 2021 https://www.R-project.org/.

7 Biedenbach DJ, Jones RN. Comparative assessment of Etest for testing susceptibilities of Neisseria gonorrhoeae to penicillin, tetracycline, ceftriaxone, cefotaxime, and ciprofloxacin: investigation using 510(k) review criteria, recommended by the Food and Drug Administration. *Journal of Clinical Microbiology* 1996; published online Dec. DOI:10.1128/jcm.34.12.3214-3217.1996.

8 Alfsnes K, Eldholm V, Olsen AO, *et al.* Genomic epidemiology and population structure of Neisseria gonorrhoeae in Norway, 2016–2017. *Microbial Genomics* 2020. DOI:10.1099/mgen.0.000359.

9 Bristow CC, Mortimer TD, Morris S, *et al.* Whole Genome Sequencing to Predict Antimicrobial Susceptibility Profiles in Neisseria gonorrhoeae. *The Journal of Infectious Diseases* 2023; : jiad027.

10 Demczuk W, Lynch T, Martin I, *et al.* Whole-Genome Phylogenomic Heterogeneity of Neisseria gonorrhoeae Isolates with Decreased Cephalosporin Susceptibility Collected in Canada between 1989 and 2013. *J Clin Microbiol* 2015; **53**: 191–200.

11 Eyre DW, De Silva D, Cole K, *et al.* WGS to predict antibiotic MICs for Neisseria gonorrhoeae. *J Antimicrob Chemother* 2017; **72**: 1937–47.

12 Ezewudo MN, Joseph SJ, Castillo-Ramirez S, *et al.* Population structure of Neisseria gonorrhoeae based on whole genome data and its relationship with antibiotic resistance. *PeerJ* 2015; **3**: e806.

13 Golparian D, Bazzo ML, Golfetto L, *et al.* Genomic epidemiology of Neisseria gonorrhoeae elucidating the gonococcal antimicrobial resistance and lineages/sublineages across Brazil, 2015–16. *Journal of Antimicrobial Chemotherapy* 2020; **75**: 3163–72.

14 Golparian D, Kittiyaowamarn R, Paopang P, *et al.* Genomic surveillance and antimicrobial resistance in Neisseria gonorrhoeae isolates in Bangkok, Thailand in 2018. *Journal of Antimicrobial Chemotherapy* 2022; : dkac158.

15 Grad YH, Harris SR, Kirkcaldy RD, *et al.* Genomic Epidemiology of Gonococcal Resistance to Extended-Spectrum Cephalosporins, Macrolides, and Fluoroquinolones in the United States, 2000–2013. *J Infect Dis* 2016; **214**: 1579–87.

16 Lan PT, Golparian D, Ringlander J, Van Hung L, Van Thuong N, Unemo M. Genomic analysis and antimicrobial resistance of Neisseria gonorrhoeae isolates from Vietnam in 2011 and 2015-16. *J Antimicrob Chemother* 2020; **75**: 1432–8.

17 Lee RS, Seemann T, Heffernan H, *et al.* Genomic epidemiology and antimicrobial resistance of Neisseria gonorrhoeae in New Zealand. *J Antimicrob Chemother* 2018; **73**: 353–64.

18 Pinto M, Borges V, Isidro J, *et al.* Neisseria gonorrhoeae clustering to reveal major European whole-genome-sequencing-based genogroups in association with antimicrobial resistance. *Microb Genom* 2020; **7**: 000481.

19 Reimche JL, Chivukula VL, Schmerer MW, *et al.* Genomic analysis of the predominant strains and antimicrobial resistance determinants within 1479 Neisseria gonorrhoeae isolates from the U.S. Gonococcal Isolate Surveillance Project in 2018. *Sex Transm Dis* 2021; published online May 14. DOI:10.1097/OLQ.0000000000001471.

20 Sánchez-Busó L, Golparian D, Corander J, *et al.* The impact of antimicrobials on gonococcal evolution. *Nature Microbiology* 2019; **4**: 1941–50.
